# Prevalence of amyloid blood clots in COVID-19 plasma

**DOI:** 10.1101/2020.07.28.20163543

**Authors:** Etheresia Pretorius, Chantelle Venter, Gert Jacobus Laubscher, Petrus Johannes Lourens, Janami Steenkamp, Douglas B Kell

**Author notes:** **Corresponding authors: Etheresia Pretorius**, Department of Physiological Sciences, Stellenbosch University, Private Bag X1 Matieland, 7602, SOUTH AFRICA, http://www.resiapretorius.net/, **Douglas B. Kell**, Department of Biochemistry and Systems Biology, Institute of Systems, Molecular and Integrative Biology, University of Liverpool, Crown St, Liverpool L69 7ZB, UK, http://dbkgroup.org/.

## Abstract

The rapid detection of COVID-19 uses genotypic testing for the presence of SARS-Cov-2 virus in nasopharyngeal swabs, but it can have a poor sensitivity. A rapid, host-based physiological test that indicated whether the individual was infected or not would be highly desirable. Coagulaopathies are a common accompaniment to COVID-19, especially micro-clots within the lungs. We show here that microclots can be detected in the native plasma of COVID-19 patient, and in particular that such clots are amyloid in nature as judged by a standard fluorogenic stain. This provides a rapid and convenient test (P<0.0001), and suggests that the early detection and prevention of such clotting could have an important role in therapy.

## Introduction

The standard method for detecting infection with SARS-CoV-2 leading to COVID-19 disease involves a genotypic (PCR) test for the virus on nasopharyngeal swabs, but it is not particularly pleasant and can have poor sensitivity (*1-4*). What would be desirable is a rapid and phenotypic test on the host that indicates the presence, and if possible the severity, of the consequences of infection. Presently, the standard method for this is based on CT chest scans for pneumonia, which have high sensitivity but lower specificity (see (*5-7*) and below), but this is neither cheap nor universally available.

It is widely recognised (*8-13*) that extensive blood clotting has a major role in the pathophysiology of COVID-19 disease severity and progression, yet so can excessive bleeding (*14, 15*). The solution to this apparent paradox lies in the recognition (*16*) that these phases are separated in time, with excessive clotting preceding the later bleeding that is mediated by the clotting-induced depletion of fibrinogen and of von Willebrand factor (VWF). This first phase is accompanied by partial fibrinolysis of the formed clots, and an extent of D-dimer formation that is predictive of clinical outcomes (*17*). These features, together with the accompanying decrease in platelets (thrombocytopaenia), leads to the subsequent bleeding. Thus it is suggested that the application of suitably monitored levels of anti-clotting agents in the earlier phase provides for a much better outcome (*10, 16*).

As well as the extent of clotting, including the life-threatening disseminated intravascular coagulation (DIC) (*12*), a second issue pertains to its nature. Some years ago, we discovered that in the presence of microbial cell wall components (*18, 19*), and in a variety of chronic, inflammatory diseases (*20-22*) (including sepsis (*23*)), blood fibrinogen can clot into an anomalous, amyloid form (*24*). These forms are easily detected by a fluorogenic stain such as thioflavin T, or the so-called Amytracker stains (*25*). In all cases, however, these experiments were performed *in vitro* using relevant plasma, with clotting being induced by the addition of thrombin. This was also the case for plasma from COVID-19 patients, but the signals were so massive that they were essentially off the scale. However, as we report here, the plasma of COVID-19 patients carries a massive load of <u>preformed</u> amyloid clots (with no thrombin being added), and this therefore provides a rapid and convenient test for COVID-19.

## Methods

### Ethical considerations

Ethical approval for blood collection and analysis of the patients with COVID-19 and healthy individuals, was given by the Health Research Ethics Committee (HREC) of Stellenbosch University (reference number: 9521). This laboratory study was carried out in strict adherence to the International Declaration of Helsinki, South African Guidelines for Good Clinical Practice and the South African Medical Research Council (SAMRC), Ethical Guidelines for research. Oral consent was obtained from all participants prior to any sample collection.

### Patient sample

#### Covid-19 patients

20 COVID-19-positive samples (11 males and 9 females) were obtained and blood samples collected before treatment was embarked upon. Blood samples were collected by JS. Platelet poor plasma (PPP) prepared and stored at -80°C, until fluorescent microscopy analysis.

#### Healthy samples

Our healthy sample was 10 age-matched controls (4 males and 6 females), previously collected and stored in our plasma repository. They were non-smokers, with CRP levels within healthy ranges, and not on any anti-inflammatory medication.

### Lung CT scans

Amongst the COVID-19 patient sample 10 patients were admitted, but stabilized and blood drawn and sent home for observation. Where patients were clinically deemed as moderate or severely ill, CT scans of the patients were performed to determine the severity of the lung pathology. We divided our sample into mild disease (no CT scan) and moderate to severely ill. The CT scan and severity score (*26*) confirmed moderate to severely ill patients according to the ‘ground glass’ opacities in the lungs.

### Fluorescent Microscopy of patient whole blood and platelet poor plasma

Fluorescent (anomalous) amyloid signals present in PPP from COVID-19 patients and healthy age-matched individuals were studied using PPP that was stored at -80°C. On the day of analysis, PPP was thawed and incubated with thioflavin T (ThT; 5 µM final concentration). Following this, the sample was incubated for 30 min (protected from light) at room temperature. PPP smears were then created by transferring a small volume (5 µl) of the stained PPP sample to a microscope slide (similar methods were followed to create a blood smear). A cover slip was placed over the prepared smear and viewed using a Zeiss AxioObserver 7 fluorescent microscope with a Plan-Apochromat 63x/1.4 Oil DIC M27 objective. Unstained samples were also prepared with both healthy and COVID-19 PPP, to assess any autofluorescence. Micrograph analysis was done using ImageJ (version 2.0.0-rc-34/1.5a). The % area of amyloid were calculated using the thresholding method. Statistical analysis was done using Graphpad, Prism 8 (version 8.4.3).

## Results

Age-matched COVID-19 (average age 49.9y) and healthy individuals (49.05y), were used in this analysis (p= 0.065). Figure 1 shows representative CT scans of four of the COVID-19 patients. Raw data is shared in https://1drv.ms/u/s!AgoCOmY3bkKHirZOu5YKPlq1×5f1AQ?e=xmWGKm

**Figure 1A to D:**
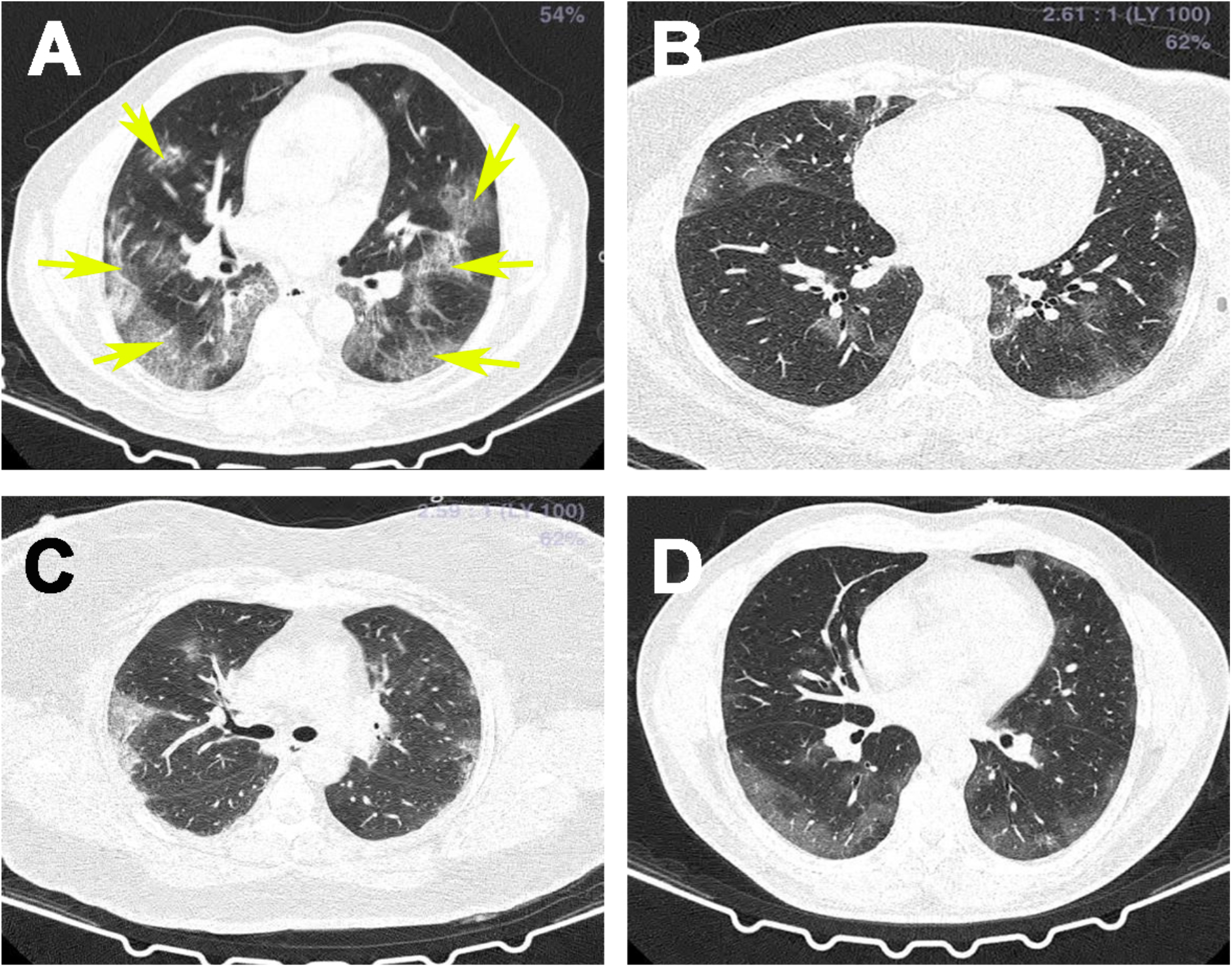
Representative CT scans of a COVID-19 patient. Yellow arrows show ground glass opacities.

Figure 2 to 4 show representative fluorescent micrographs of PPP from healthy and COVID-19 individuals. In healthy PPP smears (Figure 2), very little ThT fluorescent signal is visible, while in COVID-19 individuals (Figure 3), abundant amyloid signal is noted. Note that these signals were as received; no thrombin was added to induce clotting. Figure 4 shows the additional presence of fibrous or cellular deposits in the PPP smears. From their appearance, some of these deposits seem to have originated from endothelial cells. There have been reports of extensive endotheliopathy in COVID-19 patients (*27*). Figure 5A and B show box plots of the % area of amyloid signal calculated from representative micrographs of each individual. A nonparametric one-way ANOVA test (Kruskal-Wallis test) between all three groups showed a highly significant difference (p = <0.0001). However, a Mann-Whitney analysis between the mild and the moderate to severe COVID-19 individuals showed no significant difference (p = 0.554). Amyloid formation in plasma is therefore present in the early stages of COVID-19, when the patients are sufficiently unwell to visit the hospital and in need of stabilization.

**Figure 2A to D):**
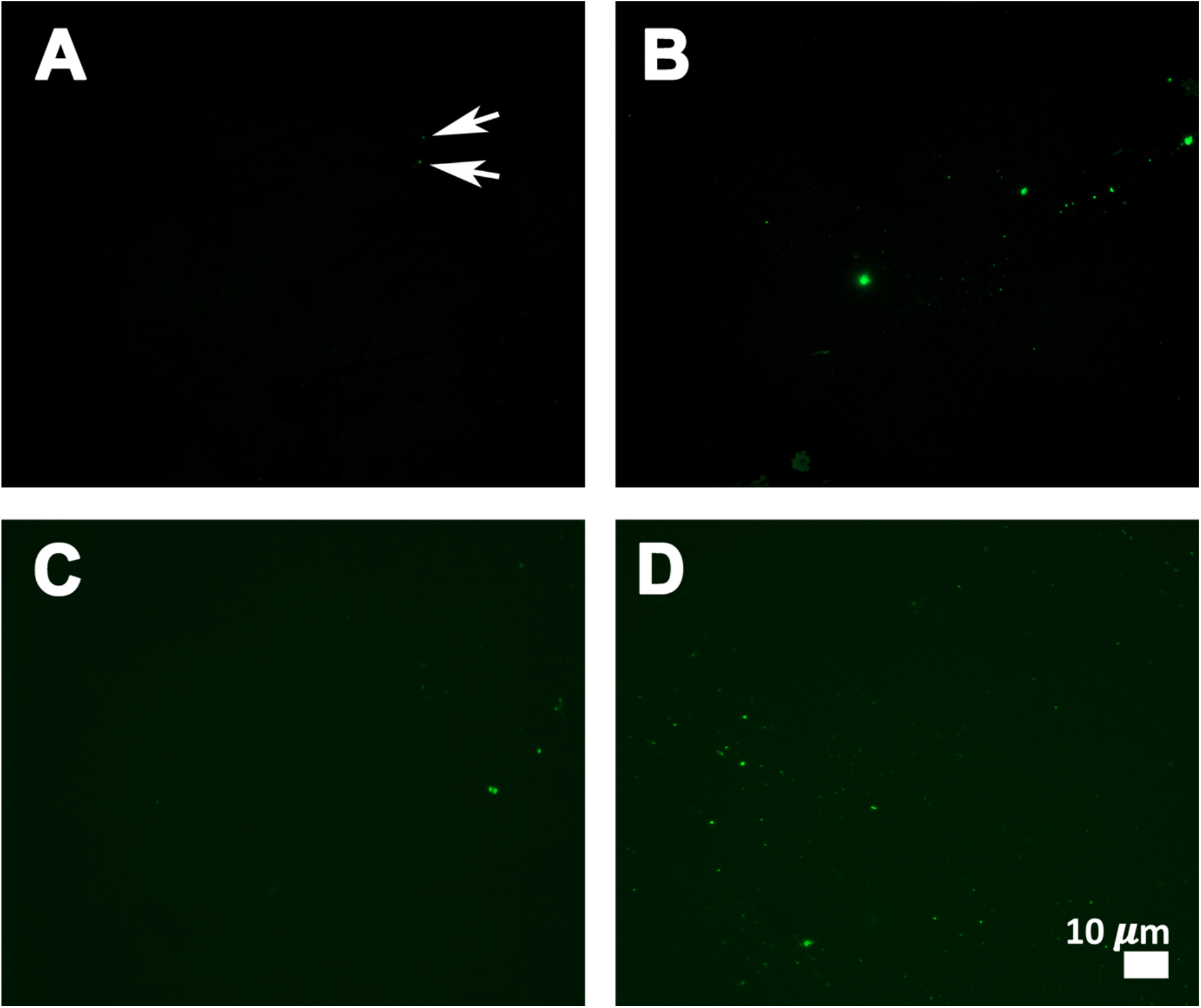
Representative fluorescent micrographs of platelet poor plasma from healthy individuals. Some signals are very slight, as shown by the arrows in **A)**.

**Figure 3A to H):**
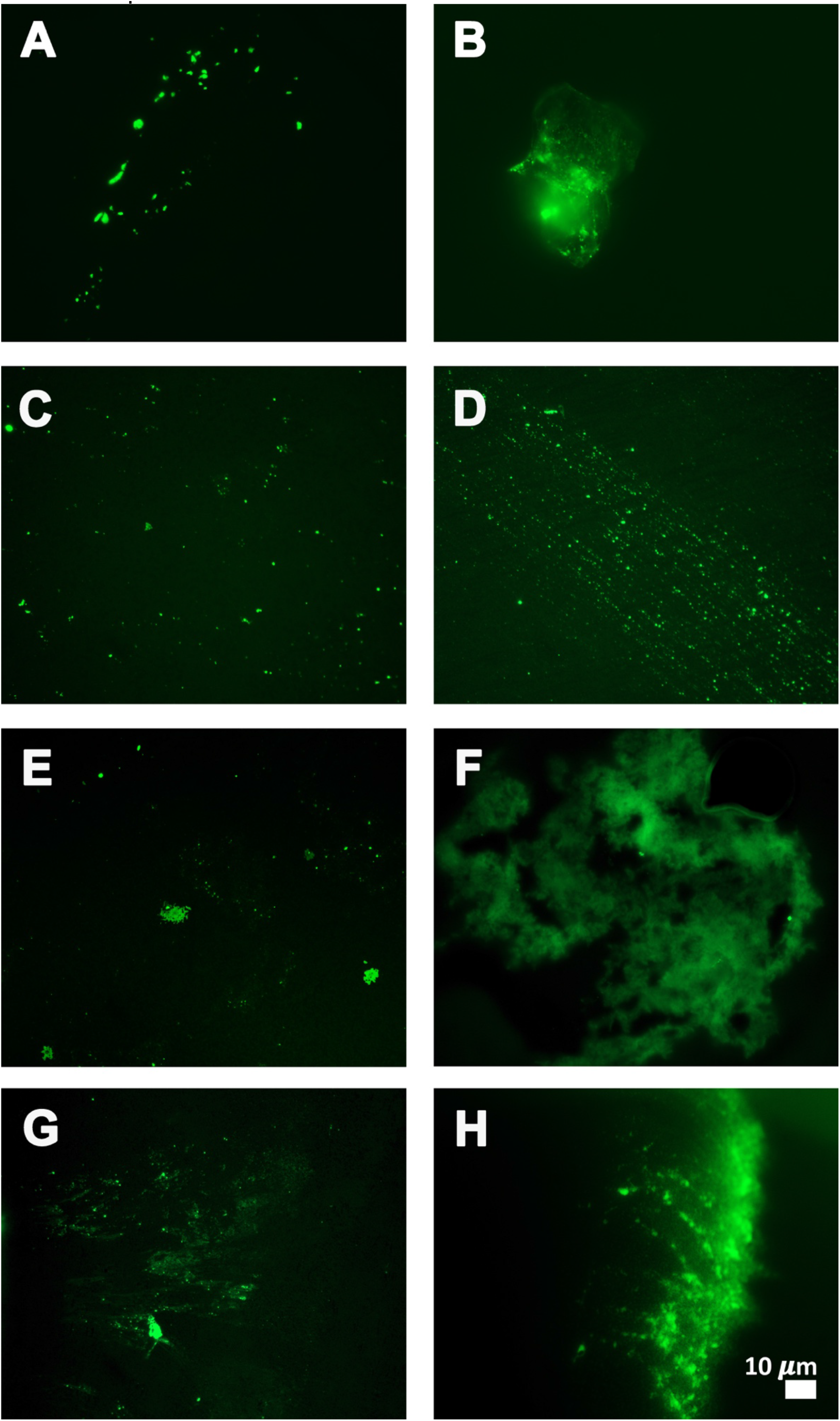
Representative fluorescent micrographs of platelet poor plasma from COVID-19 patients.

**Figure 4:**
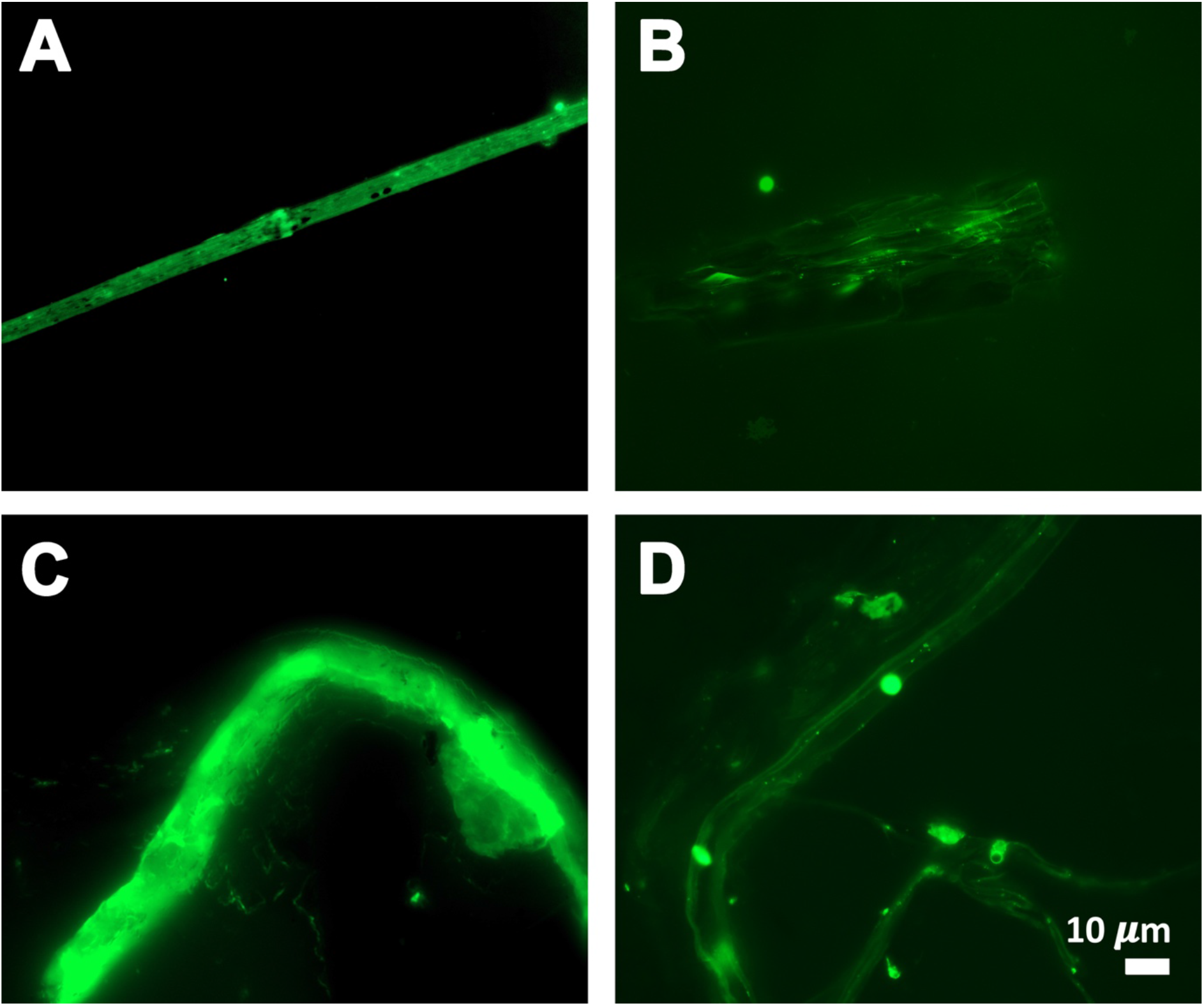
Fibrous or cellular deposits in the plasma smears of COVID-19 patients.

**Figure 5A and B:**
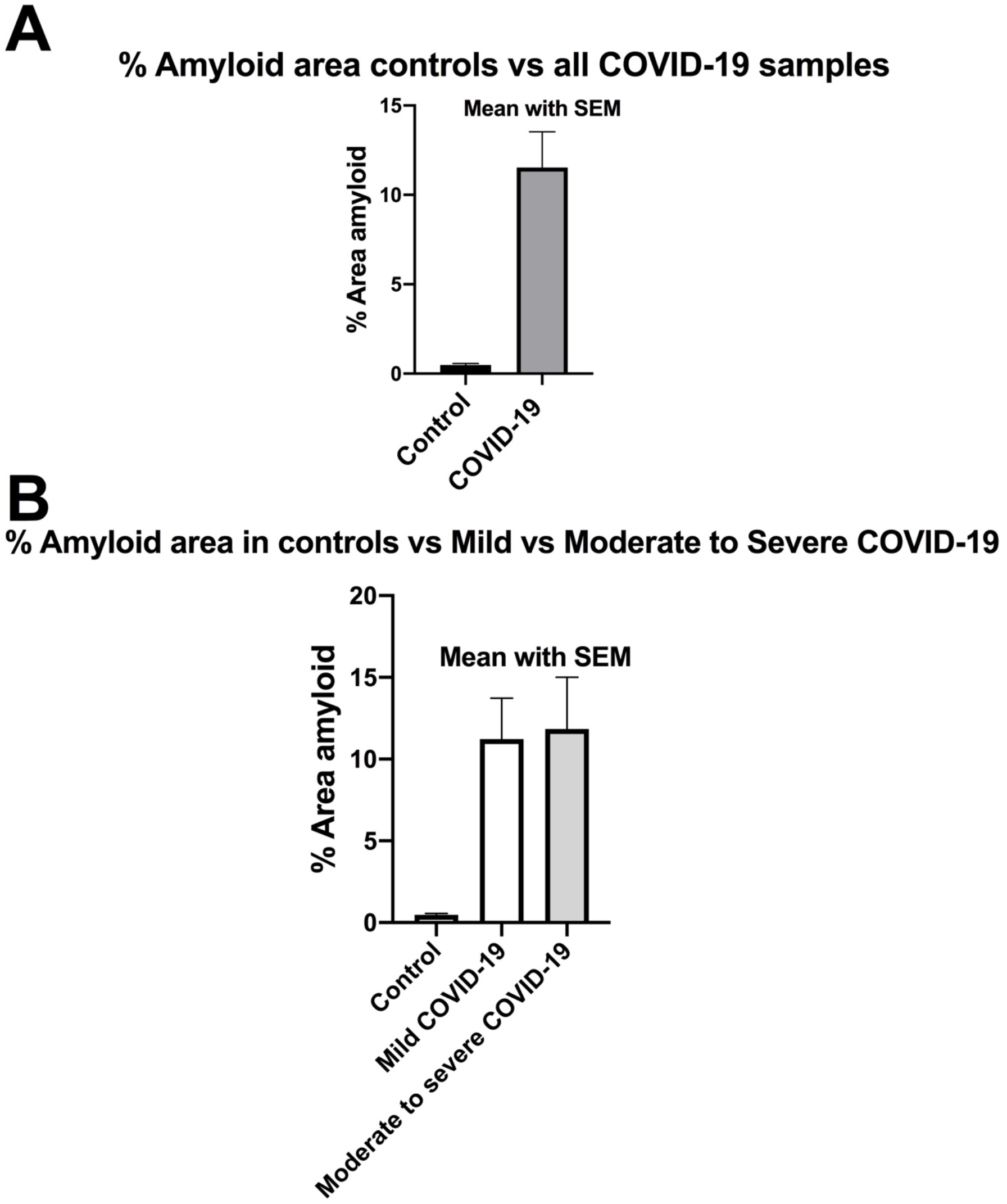
Amyloid % area in platelet poor plasma smears with mean and SEM (p = <0.0001). **A)** All controls and all COVID-19 patients. B) All controls vs 10 mild and 10 moderate to severely ill patients.

## Discussion

Strongly bound up with the coagulopathies accompanying severe COVID-19 disease is the presence of hyperferritinaemia (in cases such as the present it is a cell damage marker (*28*)) and a cytokine storm, (*29-33*) which usually occurs in the later phase of the disease (*16*). Excess iron has long been known to cause blood to clot into an anomalous form (*34*), later shown to be amyloid in nature (*18-24*). These kinds of phenomena seem to accompany essentially every kind of inflammatory disease (e.g. (*35*)), but the amyloidogenic coagulopathies are normally assessed following the *ex vivo* addition of thrombin to samples of plasma.

Many clinical features of COVID-19 are unprecedented, and here we demonstrate yet another: the presence in platelet poor plasma to which thrombin has <u>not</u> been added of amyloid microclots. This kind of phenomenon explains at once the extensive microclotting that is such a feature of COVID-19 (*8*), and adds strongly to the view that its prevention via anti-clotting agents should lie at the heart of therapy. Although fluorescence microscopy is a specialized laboratory technique, TEG is a well-known point of care technique, which is cheap and reliable. All told, the relative ease, speed (40 minutes including 30 minutes ThT incubation time) and cheapness of the assay we describe might be of considerable prognostic utility in assessing the clinical status of COVID-19 patients.

Of course this must also be monitored (e.g. via Thromboelastography (*36-39*)) lest the disease enters its later phase in which bleeding rather than clotting is the greater danger (*16*). An important consideration is that TEG can be used to study the clotting parameters of both whole blood and PPP. Whole blood TEG gives information on the clotting potential affected by the presence of both platelets and fibrinogen, while PPP TEG only presents evidence of the clotting potential of the plasma proteins (*36-39*).

Point-of-care devices and diagnostics like TEG are also particularly useful to assess fibrinolysis. In COVID-19 patients, Wright and co-workers reported fibrinolysis shutdown, confirmed by complete failure of clot lysis at 30 minutes on the TEG (*40*). Thus TEG can therefore predict thromboembolic events in patients with COVID-19 (*40*). What we have shown here is that the clotting that is commonly seen in COVID-19 patients is in an amyloid form; this alone would explain the complete shutdown of fibrinolysis and the decreased ability to pass O2 into the blood that is such a feature of the disease. Consequently, its prevention must lie at the heart of therapies.

## Data Availability

Raw data is available at: https://1drv.ms/u/s!AgoCOmY3bkKHirZOu5YKPlq1x5f1AQ?e=xmWGKm

## DECLARATIONS

### Funding

We thank the Medical Research Council of South Africa (MRC) (Self-Initiated Research Program: A0×331) for supporting this collaboration. DBK thanks the Novo Nordisk Foundation for funding (grant NNF10CC1016517).

### Competing interests

The authors declare that they have no competing interests.

### Consent for publication

All authors approved submission of the paper.

### Data sharing

https://1drv.ms/u/s!AgoCOmY3bkKHirZOu5YKPlq1×5f1AQ?e=xmWGKm

## Notes

### Competing Interest Statement

The authors have declared no competing interest.

### Clinical Trial

N/A

### Funding Statement

We thank the Medical Research Council of South Africa (MRC) (Self-Initiated Research Program: A0X331) for supporting this collaboration. DBK thanks the Novo Nordisk Foundation for funding (grant NNF10CC1016517).

### Author Declarations

Ethical approval for blood collection and analysis of the patients with COVID-19 and healthy individuals, was given by the Health Research Ethics Committee (HREC) of Stellenbosch University (reference number: 9521).

